# Alzheimer’s disease and neurodegeneration in symptomatic cerebral small vessel disease

**DOI:** 10.1101/2024.05.20.24307607

**Authors:** Philipp Arndt, Malte Pfister, Valentina Perosa, Hendrik Mattern, Jose Bernal, Anna-Charlotte John, Marc Dörner, Patrick Müller, Rüdiger C. Braun-Dullaeus, Cornelia Garz, Christopher Nelke, Alma Kokott, Robin Jansen, Michael Gliem, Sven G. Meuth, Solveig Henneicke, Stefan Vielhaber, Katja Neumann, Stefanie Schreiber

## Abstract

**Background:** Co-occurrence of cerebral small vessel disease (CSVD) is common in aging and Alzheimer’s disease (AD) dementia, but, in symptomatic CSVD prevalence and role of AD and neurodegenerative co-pathologies have been less explored.

**Methods:** *In vivo* determination of prevalence, predictors and relevance for cognition of AD and neurodegenerative co-pathologies in symptomatic CSVD, including deep perforator arteriopathy (DPA) and cerebral amyloid angiopathy (CAA), utilizing cerebrospinal fluid (CSF) biomarkers. Cross-sectional study from October 2010 to September 2021 of participants with magnetic resonance imaging (MRI) and CSF biomarkers (amyloid-β 42/40 ratio, phosphorylated-tau, total-tau, neurofilament light). Biomarker levels were compared among groups; prevalence of ATN classification subtypes was estimated and related to clinical phenotype, CSVD MRI markers and global cognition.

**Results:** The study comprised 229 individuals (median age 74 years; 47% females), 70 with AD dementia, 79 with probable CAA, 62 with DPA patients and 18 healthy controls. Participants were categorized based on the ATN classification: normal biomarkers (A-T-N-), AD pathology continuum (A+T±N±), and non-AD pathological changes, including primary age-related tauopathy (PART, A-T+N+) or isolated neurodegeneration (A-T-N+). Of 141 CSVD patients, 43 (30%) were A-T-N-, 39 (28%) A+T±N±, with lower prevalence in DPA than CAA (15% vs. 38%, p = .003), 18 (13%) A-T+N+, and 41 (29%) A-T-N+, with higher prevalence in DPA than CAA (42% vs. 19%, p = .002). A+T±N± was associated with increasing age, female sex, lobar hemorrhages and low burden of deep white matter hyperintensities and lacunes. A-T-N+ was related to younger age, symptomatic stroke and lacunes. A-T+N+ had no specific predictors, except advanced age. Each pathological ATN profile was independently related to lower Mini Mental State Examination scores (A+T±N±: B = -2.7, p = .013; A-T+N+: B = -4.6, p = .002; A-T-N+: B = -2.3, p = .034), accounting for demographics, clinical phenotype and MRI CSVD severity.

**Conclusions:** Using biomarkers, this study confirms *in vivo* that CSVD frequently co-occurs with AD or neurodegenerative pathologies, exerting independent effects on cognitive health. As disease-modifying therapies emerge, integrating interacting biomarkers will be crucial for the selection of patients with the greatest benefit.

## Background

*Postmortem* neuropathologic studies revealed that the co-occurrence of cerebral small vessel disease (CSVD) and Alzheimer’s disease (AD) or neurodegenerative pathology is common and accelerates cognitive impairment, compared to isolated pathologies [1–4].

In clinical practice, the characterization of concurrent brain pathologies *in vivo* remains rare, as typically, primary symptomatic pathologies receive focus in care. CSVD diagnosis relies on magnetic resonance imaging (MRI) detection of non-hemorrhagic and hemorrhagic lesions, with heterogeneous clinical presentation and a lack of disease-specific biofluid markers [5].Conversely, AD dementia diagnosis is based on disease-specific biofluid markers for amyloid-beta (Aβ) and phosphorylated-tau (pTau), alongside a specific cognitive profile [6]. Integrating MRI-based CSVD detection with the biomarker-driven scheme for AD pathology diagnosis, the ATN classification, facilitates identification of individuals with isolated or mixed brain pathologies.

*In vivo* studies have confirmed the significance of concurrent CSVD in AD dementia patients. Compared to cognitively unimpaired controls, they demonstrated more frequently CSVD lesions on MRI, which accelerated cognitive decline [7–12].

In contrast, less is known about mixed pathologies in patients with symptomatic CSVD. Some studies investigated concurrent Aβ or tau pathology using positron electron tomography (PET) imaging [13–16], but these studies comprise small sample sizes or lack precise clinical and neuroradiological characterization of CSVD etiology. Further, the entire ATN classification has not been fully applied in symptomatic CSVD to investigate AD (A+T±N±) and neurodegenerative non-AD (A-T+N+, A-T-N+) co-pathologies.

We aimed to overcome these knowledge gaps by applying MRI and fluid biomarkers to a clinical cohort of 229 participants, including patients with symptomatic deep perforator arteriopathy (DPA) and cerebral amyloid angiopathy (CAA), to investigate prevalence of AD and neurodegenerative non-AD co-pathologies, their predictors and relevance on global cognition *in vivo*.

## Methods

### Study population

We included 211 patients with DPA, CAA or AD dementia and 18 controls with non-specific complaints who underwent MRI and lumbar puncture (LP) for diagnostic workup between 10/2010 and 09/2021 in the Department of Neurology, Otto-von-Guericke University Magdeburg. DPA diagnosis was based on the existence of deep cerebral microbleeds (CMB), probable CAA on the Boston criteria version 2.0 [17] and AD dementia on the NINDCS/ADRDA and ATN criteria, i.e. a positive cerebrospinal fluid (CSF) AD pathology profile (A+T+N±) [18, 19]. None of the CSVD patients fulfilled the diagnostic criteria for AD dementia and all controls were free of CMB.

### Clinical data and neuropsychological assessment

Patients were characterized with regard to clinical symptomatic phenotype (cognitive impairment, symptomatic stroke, gait disturbances, seizure or a mixture of them), demographics (age, sex), years of education (available for n = 159, 69%), apolipoprotein E (ApoE) genotype (available for n = 105, 46%) and presence of vascular risk factors (available for n = 218, 95%). Global cognition was determined through Mini Mental State Examination (MMSE, available for n = 132, 58%) (for details see **Supplementary material**).

### Cerebrospinal fluid

CSF biomarker levels of AD pathology (Aβ_42/40_ ratio, pTau) and neurodegeneration (total-tau (tTau) and Neurofilament light (NfL, available for n = 136, 59%)) were used to categorize participants as A-T-N-(normal biomarker status), A+T±N± (AD pathology continuum), A-T+N+ (age-related tauopathy, PART) or A-T-N+ (isolated neurodegeneration). None of the participants was classified A-T+N- (for details see **Supplementary material**).

### MRI acquisition and analysis

For analysis, clinical 3T MRI (n = 92, 40%) or 1.5T MRI (n = 137, 60) was available and intracerebral hemorrhage (ICH), CMB, white matter hyperintensities (WMH) and lacunes were quantified according to the CSVD Standards for Reporting Vascular Changes on Neuroimaging (STRIVE) criteria [5] by one trained investigator (M.P.) blinded to demographic and clinical data. Global CSVD burden was determined through an ordinal sum score ranging from 0 to 5 points, in which one point was allocated for the presence of lacunes, 1-4 CMB and moderate WMH (WMH grade 3-4 according to Fazekas [20]) and two points were allocated for ≥5 CMB and severe WMH (WMH grade 5-6), respectively (for details see **Supplementary material**).

### Statistical analysis

Results of continuous variables were expressed as median (IQR) or mean (SD), as appropriate; results of categorical variables were expressed as proportions. Intergroup comparisons were performed in univariate analyses, using the χ^2^-test, 2-sample t-test, Mann-Whitney U-test, Kruskal-Wallis-test or ANOVA as appropriate. Pairwise post-hoc comparisons were conducted applying Kruskal-Wallis-test with Dunn’s correction and χ^2^-test or ANOVA with Bonferroni’s correction for multiple testing. Univariate tests were used to compare clinical and neuroimaging data between patients with and those without each pathological ATN profile, i.e. separately for A+T±N±, A-T+N+ and A-T-N+. To explore independent, relevant predictors of pathological ATN profiles in CSVD, we performed multivariable logistic regression analysis in a stepwise, forward-elimination approach (minimal adjusted model), which was based on the results from univariable analyses (including variables with p < .05). Linear regression was conducted to examine associations between pathological ATN profiles and MMSE score in CSVD, accounting for age, clinical phenotype and global MRI CSVD severity. (Adjusted) Significance level was set at 0.05 for all analyses. IBM SPSS Statistics 24.0 software was used for all analyses.

## Results

We included 229 participants with a median age of 74 years (IQR 67 – 79), of whom 107 (47%) were female. Diagnostic groups were distributed as follows: n = 18 controls and n = 211 patients with DPA (n = 62, 27%), probable CAA (n = 79, 34%) or AD dementia (n = 70, 31%). Out of all AD dementia patients, n = 12 (17%) had CMB, of whom n = 10 met the Boston criteria 2.0 for concurrent probable CAA and n = 2 had concurrent DPA. Key characteristics of the cohort and statistical group comparisons are shown in **Table 1 and Supplementary Table 1**.

**Table 1.** Inter-group comparison of patient characteristics.

|  | CON<br>n = 18 | DPA<br>n = 62 | CAA<br>n = 79 | AD<br>dementia<br>n = 70 | Univariate<br>analysis | Significant pairwise<br>post-hoc tests |
| --- | --- | --- | --- | --- | --- | --- |
| **Demographics** |  |  |  |  |  |  |
| Age [years] | 65<br>(57-73) | 70<br>(61-78) | 75<br>(70-80) | 75<br>(71-79) | H = 23.36<br>**p < .001** | CON vs. CAA, AD: **p = .001**<br>DPA vs. CAA, AD: **p < .040** |
| Female sex | 3 (17%) | 27 (44%) | 34 (43%) | 43 (61%) | $\chi^2 = 13.30$<br>**p = .004** | CON vs. AD: **p = .006** |
| Education<br>[years]<sup>a</sup> | 14<br>(13-16) | 13<br>(11-16) | 13<br>(11-16) | 13<br>(13-15) | H = 5.14<br>p = .162 |  |
| **Clinical phenotype** |  |  |  |  |  |  |
| Cognitive<br>impairment | 0 (0%) | 28 (45%) | 52 (66%) | 70 (100%) | $\chi^2 = 63.57$<br>**p < .001** | CON vs. DPA, CAA: **p < .002**<br>CON, DPA, CAA vs. AD: **p < .001** |
| Previous<br>symptomatic<br>stroke | 0 (0%) | 26 (42%) | 29 (37%) | 2 (3%) | $\chi^2 = 39.68$<br>**p < .001** | CON vs. DPA, CAA: **p < .020**<br>DPA, CAA vs. AD: **p < .001** |
| Gait<br>disturbances | 0 (0%) | 17 (27%) | 17 (22%) | 13 (19%) | $\chi^2 = 6.67$<br>p = .083 | |
| Previous<br>seizure | 0 (0%) | 15 (24%) | 15 (19%) | 2 (3%) | $\chi^2 = 17.16$<br>**p = .001** | DPA, CAA vs. AD: **p < .02** |
| **Risk factors** |  |  |  |  |  |  |
| ApoE4<br>positive | n.a. | 11/31<br>(36%) | 13/35<br>(37%) | 26/39<br>(67%) | $\chi^2 = 9.04$<br>**p = .011** | |
| Hypertension | 10/18<br>(56%) | 56/60<br>(93%) | 63/72<br>(88%) | 53/68<br>(78%) | $\chi^2 = 16.76$<br>**p < .001** | CON vs. DPA, CAA: **p < .02** |
| Dyslipidemia | 5/18<br>(28%) | 30/60<br>(50%) | 27/72<br>(38%) | 22/68<br>(32%) | $\chi^2 = 5.34$<br>p = .149 | |
| Type 2<br>diabetes | 1/18<br>(6%) | 13/60<br>(22%) | 20/72<br>(28%) | 21/68<br>(31%) | $\chi^2 = 5.48$<br>p = .139 | |
| **Cognition** |  |  |  |  |  |  |
| MMSE<sup>b</sup> |  | 24.8 (4.0) | 22.8 (5.0) | 19.5 (6.4) | F = 9.60<br>**p < .001** | DPA, CAA vs. AD: **p < .02** |

Data are represented as median (IQR), mean (SD) or n (%). Significant p-values are marked in bold.
<sup>a</sup> available for n = 159 (69%), <sup>b</sup> available for n = 132 (58%).
Abbreviations: AD, Alzheimer's disease; ApoE, apolipoprotein E; CAA, cerebral amyloid angiopathy; CON, controls; DPA, deep perforator arteriopathy; MMSE, Mini Mental State Examination; n.a., not available.

### One quarter of CSVD patients is within the AD pathology continuum

Of 141 CSVD patients, 39 (28%) were within the AD pathology continuum (A+T±N±) according to ATN, with lower prevalence in DPA than CAA (15% vs. 38%, p = .003), particularly for A+T+N+ (3% vs. 27%, p < .001) (**Table 2**). **Figure 1** and **Supplementary Table 2** demonstrate the distribution of the remaining ATN profiles within the AD pathology continuum across the cohort. In CSVD, continuous AD pathology biomarker levels were in between those of controls and AD dementia, with lower AD pathology (higher Aβ_42/40_ ratio, lower pTau) in DPA than CAA (**Supplementary Figure 1** and **Supplementary Table 3**).

**Figure 1.**
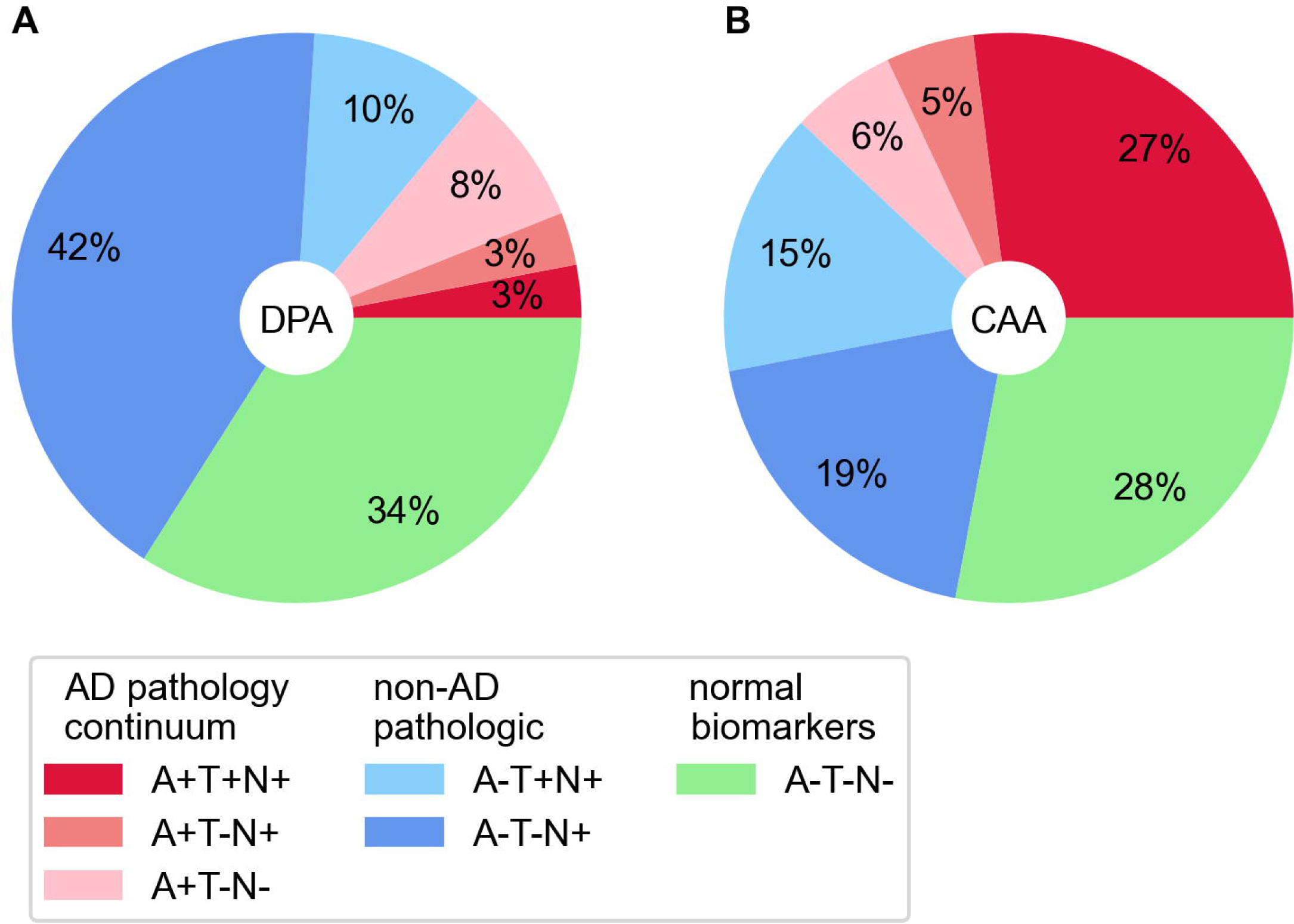
Relative frequencies of ATN profiles in patients with DPA and CAA. None of the participants were A+T+N- or A-T+N-. Abbreviations: A, amyloid-beta; AD, Alzheimer’s disease; CAA, cerebral amyloid angiopathy; DPA, deep perforator arteriopathy; T, phosphorylated Tau; N, neurodegeneration.

**Table 2.** Inter-group comparison of ATN profiles and neurodegeneration markers.

|  | CON<br>n = 18 | DPA<br>n = 62 | CAA<br>n = 79 | AD<br>dementia<br>n = 70 | Univariate<br>analysis | Significant pairwise<br>post-hoc tests |
| --- | --- | --- | --- | --- | --- | --- |
| **Pathological ATN classification profiles** |  |  |  |  |  |  |
| A+T±N± | | 9 (15%) | 30 (38%) | | $\chi^2 = 8.93$<br>**$p = .003$** | |
| A-T+N+ | | 6 (10%) | 12 (15%) | | $\chi^2 = 0.98$<br>$p = .323$ | |
| A-T-N+ | | 26 (42%) | 15 (19%) | | $\chi^2 = 10.02$<br>**$p = .002$** | |
| **Neurodegeneration markers** |  |  |  |  |  |  |
| tTau-positive | 0<br>(0%) | 24<br>(39%) | 46<br>(58%) | 65<br>(93%) | $\chi^2 = 65.46$<br>**$p < .001$** | CON vs. DPA, CAA: **$p < .01$**<br>AD vs. CON, DPA, CAA: **$p < .001$** |
| NfL-positive | 0/8<br>(0%) | 23/41<br>(56%) | 22/42<br>(52%) | 3/45<br>(7%) | $\chi^2 = 33.65$<br>**$p < .001$** | CON vs. DPA, CAA: **$p < .05$**<br>AD vs. DPA, CAA: **$p < .001$** |

Data are represented as median (IQR), mean (SD) or n (%). Significant p-values are marked in bold.
<sup>a</sup> available for n = 136 (59%).
Abbreviations: AD, Alzheimer's disease; CAA, cerebral amyloid angiopathy; CON, controls; CSF, cerebrospinal fluid; DPA, Deep perforator arteriopathy; NfL, neurofilament light chain; tTau, total-tau.

### Neurodegeneration is frequent and commonly related to non-AD pathological change in CSVD

Of 141 CSVD patients, 88 (62%) were N+, of whom 59 (42%) were attributable to non-AD pathological ATN profiles, including 18 (13%) with A-T+N+ and 41 (29%) with A-T-N+. Prevalence of A-T-N+ was higher in DPA than CAA (42% vs. 19%, p = .002). Prevalence of NfL positivity was significantly higher in DPA and CAA, compared to AD dementia (56% and 52% vs. 7%), whereas prevalence of tTau positivity was lower in DPA and CAA compared AD dementia (39% and 58% vs. 93%) (**Figure 1**, **Table 2**). Continuous neurodegeneration biomarker levels are demonstrated in **Supplementary Figure 1** and **Supplementary Table 3.**

### MRI CSVD pathology and clinical phenotype are associated with pathological ATN profiles

We next investigated the relationship between pathological ATN profiles, MRI CSVD pathology and clinical phenotypes in CSVD. Significant variables (p < .05) of univariate comparisons between patients with and without pathological ATN profiles (separately for A+T±N±, A-T+N+, A-T-N+ in **Supplementary Table 4**) were included in logistic regression analysis to explore independent predictors. A+T±N± was independently associated with lobar ICH (odds ratio (OR) 3.19 [95% confidence interval (95% CI) 1.14, 8.95], p = .028), female sex (OR 2.54 [95% CI 1.01, 6.47], p = .049), increasing age (per increase of 5 years: OR 1.75 [95% CI 1.25, 2.44], p < .001) and higher numbers of lobar CMB (per increase of 5 counts: OR 1.13 [95% CI 1.03, 1.24], p = .007). Contrary, deep WMH (OR 0.53 [95% CI 0.31, 0.92], p = .024) and deep lacunes (OR 0.16 [95% CI 0.03, 0.79], p = .024) decreased relative A+ T±N± probability. A-T-N+ was independently related to previous symptomatic stroke (OR 3.39 [95% CI 1.50, 7.64], p = .003) and numbers of lacunes (OR 1.16 [95% CI 1.03, 1.30], p = .012), whereas higher age decreased the relative A-T-N+ probability (per increase of 5 years: OR 0.76 [95% CI 0.60, 0.95], p = .011). A-T+N+ had no specific predictors except increasing age (per increase of 5 years: OR 1.37 [95% CI 1.00, 1.88], p = .048) (**Table 3**). The predictive effects of age and female sex for A+T±N± were driven by DPA, while the effects of lobar ICH and CMB were driven by CAA (**Supplementary Table 5**).

**Table 3.** Multivariable logistic regression analyses of associations with pathological ATN profiles in CSVD patients.

| ATN profile | Independent variable | OR (95% CI) | P-value | Model |
| --- | --- | --- | --- | --- |
| A+T±N± | Presence of lobar ICH | 3.19 (1.14, 8.95) | **.028** | **p < .001**, f<sup>2</sup> = .72 |
|  | Female sex | 2.54 (1.01, 6.47) | **.049** |  |
|  | Age | 1.75 (1.25, 2.44) | **< .001** |  |
|  | Number of lobar CMB | 1.13 (1.03, 1.24) | **.007** |  |
|  | Deep WMH score | 0.53 (0.31, 0.92) | **.024** |  |
|  | Presence of deep lacune | 0.16 (0.03, 0.79) | **.024** |  |
| A-T+N+ | Age | 1.37 (1.00, 1.88) | **.048** | **p = .049**, f<sup>2</sup> = .06 |
| A-T-N+ | Previous stroke | 3.39 (1.50, 7.64) | **.003** | **p < .001**, f<sup>2</sup> = .34 |
|  | Number of lacunes | 1.16 (1.03, 1.30) | **.012** |  |
|  | Age | 0.76 (0.60, 0.95) | **.011** |  |

Odds ratios with 95% confidence intervals are related to an increase of 5 years for age and 5 counts for number of lobar CMB. Significant p-values are marked in bold.
Abbreviations: CI, confidence interval; CMB, cerebral microbleeds; CSVD, cerebral small vessel disease; ICH, intracerebral hemorrhage; WMH, white matter hyperintensities.

### AD and non-AD pathological ATN profiles negatively impact global cognition in CSVD

In a subset of CSVD patients with available MMSE (n = 79, 54%; n = 36 DPA, n = 43 CAA), pathological ATN profiles were related to global cognition using linear regression analyses accounting for age, clinical phenotype and MRI global CSVD severity. The MMSE score was independently associated with all three distinct pathological ATN profiles with decreasing effect size in the following order: A-T+N+ (B = -4.6 [-7.4, -1.8], p = .002), A+T±N± (B = -2.7 [-4.8, -0.6], p = .013) and A-T-N+ (B = -2.3 [-4.3, -0.2], p = .034) (**Table 4)**.

**Table 4.** Associations of pathological ATN profiles with global cognition in CSVD patients.

| Dependent variable | Independent variables | B (95% CI) | $\beta$ | P-value | Model |
| --- | --- | --- | --- | --- | --- |
| MMSE | A+T±N± | -2.7 (-4.8, -0.6) | -0.27 | **.013** | **p < .001**<br>$R^2_{adj} = .54$ |
|  | A-T+N+ | -4.6 (-7.4, -1.8) | -0.30 | **.002** |  |
|  | A-T-N+ | -2.3 (-4.3, -0.2) | -0.22 | **.034** |  |

Multivariate linear regression analyses with pathological ATN profiles as independent variables and MMSE (available for $n = 74$ CSVD patients, 53%; $n = 36$ DPA, $n = 43$ CAA) as the dependent variable were conducted. The model included age, clinical phenotype (cognitive impairment, previous symptomatic stroke, gait disturbances, seizure) and a CSVD severity sum score as confounding variables. Significant p-values are marked in bold.
Abbreviations: CI, confidence interval; CSVD, cerebral small vessel disease; MMSE, Mini Mental State Examination.

## Discussion

AD pathology and neurodegeneration CSF biomarkers were investigated in symptomatic CSVD patients. AD pathology continuum (A+T±N±) was found in nearly one third of patients, and was associated with older age, female sex, lobar hemorrhages and fewer deep WMH and lacunes. Non-AD pathological changes became obvious in 42%, with isolated neurodegeneration (A-T-N+), in particularly younger patients with symptomatic stroke or lacunes, or PART (A-T+N+), which was solely predicted by advanced age. All three pathological ATN profiles were independently associated with lower global cognition, regardless of clinical CSVD phenotype. Using CSF biomarkers, we confirm *in vivo* that CSVD and AD or non-AD neurodegenerative pathologies co-occur, characterizing different pathological subtypes of symptomatic CSVD with relevance for cognitive health.

### AD pathology continuum in CSVD

Based on the CSF Aβ_42/40_ ratio we report positivity rates of 15% (DPA) to 38% (CAA), while rates of others were 9% to 32% in DPA [13, 15, 21, 22] or 44% to 75% in CAA [13, 14, 16, 22, 23]. Differences in CAA might derive from methodological disparities. First, prior studies included clinical AD dementia with concurrent CAA. In our cohort, 10 out of 70 (14%) AD dementia patients met the Boston criteria v2.0 and would have increased A+T±N± rates within our CAA sample from 38% to 45%. Conversely, none of our CAA patients met the diagnostic criteria for AD dementia (NINDCS/ADRDA, ATN criteria). Next, we applied the updated Boston criteria v2.0 for patient selection, which display superior accuracy compared to v1.5 for the detection of early CAA, which is presumably characterized by lower A+T±N± rates [24]. Comparing the Boston criteria v2.0 vs. v1.5, we confirmed lower A+T±N± rates in v2.0, while other pathological ATN profiles remained unchanged (**Supplementary Table 6**).

There is scarce knowledge about the stages of the AD pathology continuum, i.e. A+T-vs. A+T+, in DPA or CAA. We show that the majority of A+ DPA patients displayed isolated Aβ pathology without pTau positivity, while the majority of A+ CAA patients were already in advanced stages with fully developed AD pathology (A+T+). These findings might mirror the fact that DPA and AD pathology develop additively with aging, while CAA and AD pathology are mechanistically interlinked promoting faster progression of amyloid and tau accumulation. This is supported by the fact, that in the current study the association between age and AD pathology was mainly driven by DPA patients. Likewise, neuropathological data from early and late onset AD dementia showed that the presence of DPA co-pathologies (lacunar or small infarcts) were associated with advanced age, while CAA co-pathology developed independently from age.[25] Pathophysiologically, CAA results from impaired clearance of soluble Aβ from the brain, which accelerates parenchymal Aβ deposition and, hence, tau accumulation [26]. *Vice versa* AD pathology promotes clearance demands and, thus, CAA, overall explaining the relationship [24, 27].

The mechanistic role of DPA pathologies such as arteriolosclerosis in cognitive decline is still a matter of debate. A neuropathological study has recently suggested that arteriolosclerosis impairs cognitive health mainly through the development of infarcts rather than through the progression of AD pathology [27]. Here, deep lesions such as WMH and lacunes that reflect DPA downstream pathologies were associated with a lower relative AD pathology risk. This is in line with recent MRI studies, where lacunes and WMH were associated with the absence of Aβ co-pathology in cognitively impaired CSVD patients [23, 28].

CAA explained the association between lobar hemorrhages and AD pathology. In CAA, lobar hemorrhages reflect advanced disease stages, where chronic vascular Aβ exposure induces smooth muscle cell loss, creating a rupture-prone vascular segment [24]. A study in CAA patients has similarly associated frequent lobar CMB with the AD pathology continuum [23] and, further, a more precise anatomical location of lobar CMB in strictly juxta- and intracortical areas was associated with Aβ accumulation in spontaneous ICH [29]. In DPA, lobar hemorrhages reflect advanced hypertensive CSVD pathology as well (but not CAA) [30], which may explain the absent association with Aβ co-pathology.

### Non-AD pathological changes

Suspected non-AD pathological changes refers to individuals with normal Aβ, but abnormal neurodegeneration biomarkers (A-T-N+, A-T+N+) [31]. These profiles have not yet been studied in CSVD. Notably, N+ was found in >60% of CSVD patients, mainly attributable to non-AD pathological changes.

N+ was based on abnormal CSF tTau and NfL, both released into the extracellular space upon neuronal injury or death. CSF NfL showed no sensitivity to AD dementia but was strongly increased in CSVD patients, suggesting it as a promising biofluid neurodegeneration marker that distinguishes between AD and CSVD pathology. The absent correlation between CSF NfL and AD pathological markers in CSVD support this assumption (data not shown), as well as recent studies that have related NfL, WMH severity and cognitive decline in CSVD [32–38].

The A-T+N+ biomarker profile is indicative of neurodegenerative fibrillary tau tangle pathology without Aβ accumulation and might indicate PART. PART is common among the elderly and can lead to cognitive impairment. The strongest neuropathological predictor for cognitive impairment in PART is CSVD co-pathology [39, 40]. Our results showed that aging was the only predictor of an A-T+N+ profile, which suggests CSVD and PART rather develop additively with aging. While the effects of vascular Aβ are better understood, it remains uncertain how pathological tau interacts with vascular pathology. Upcoming evidence suggests that in absence of Aβ, tau induces morphological changes in blood vessels, which impairs blood flow and accelerates neurodegeneration [41]. However, further research is needed to understand how vascular pathology interacts together with tau accumulation to accelerates cognitive decline.

### Cognition

While different pathological ATN profiles appear to characterize different CSVD subtypes, they similarly relate to poor global cognition. Additive Aβ co-pathology has been associated with accelerated longitudinal cognitive decline in lacunar stroke, CAA and CSVD with preexisting cognitive impairment [16, 21, 42]. One may speculate that this particular CSVD cohort could also benefit from Aβ modifying immunotherapy. Considerations and according studies have to be taken against the background that therapeutic mechanisms rely on vascular clearance, which might be impaired in CSVD, and related side effects, i.e., amyloid-related imaging abnormalities, have to be monitored even more carefully. There are no studies on the cognitive outcome of non-AD pathological ATN profiles in CSVD, although NfL alone has been associated with cognitive decline (as explained above). Similar to AD dementia, our study confirms that neurodegeneration leads to cognitive decline regardless of etiology.

### Strengths and limitations

The main strength of our study is the use of large and clinically diverse CSVD cohort along the whole spectrum of different disease stages, which had complete information on ATN profiles and STRIVE. The approach is of translational value, as the ATN classification is easy to implement and provides bed-side information, along with the separate consideration of DPA and CAA, which have different disease trajectories and therapies. Further, the CSF Aβ_42/40_ ratio, used here, displays superior accuracy compared to CSF Aβ42 alone in reference to neuropathological Aβ burden and does not correlate with neuropathological CAA severity, making it an attractive biomarker to study (parenchymal) Aβ co-pathology *in vivo* [43].

Our study faces limitations. There were missing data for vascular risk factors, ApoE status, CSF NfL and MMSE, which is based on the clinical nature of the cohort. We cannot rule out that additional age-related neurodegenerative co-pathologies (e.g. alphasynucleinopathies) have contributed to non-AD pathological changes [44]. Notably, differences of the prevalence of ATN subtypes can derive from different locally established cut-offs, and our results should thus be validated in a multicenter approach.

## Conclusions

The development of multiple interacting brain pathologies during aging and their mutual relevance for brain health requires multiple biomarkers to better understand the complexity of brain pathologies *in vivo*. Further work is necessary to fully understand the interplay of vascular and neurodegenerative diseases and capture the value of co-pathology testing. Particularly with the emergence of disease-modifying therapies for neurodegenerative and cerebrovascular diseases it will be necessary to integrate interacting biomarkers for the selection of patients with greatest benefit.

## Data Availability

The datasets analyzed during the current study are available from the corresponding author on reasonable request.

## List of abbreviations

Aβ: amyloid-beta
AD: Alzheimer’s disease
ApoE: apolipoprotein E
CAA: cerebral amyloid angiopathy
CI: confidence interval
CMB: cerebral microbleeds
CON: controls
CSF: cerebrospinal fluid
CSVD: cerebral small vessel disease
DPA: deep perforator arteriopathy
ICH: intracerebral hemorrhage
LP: lumbar puncture
MMSE: Mini Mental State Examination
MRI: magnetic resonance imaging
NfL: Neurofilament light
PART: primary age-related tauopathy
PET: positron electron tomography
pTau: phosphorylated-tau
STRIVE: Standards for Reporting Vascular Changes on Neuroimaging
tTau: total-tau
WMH: white matter hyperintensities.

## Declarations

### Ethics approval and consent to participate

The local ethic committee (Ethikkommission, Otto-von-Guericke-Universität Magdeburg; No. 07/17, addendum 11/2021) approved this retrospective study.

### Consent for publication

Informed consent was obtained from all participants for anonymized retrospective analysis of their data conducted for clinical diagnostics.

### Authors’ contributions

P.A., M.P., K.N. and S.S. have contributed to the conception or design of the work. P.A., M.P., A.C.J., C.G., K.N. and S.V. have contributed to the acquisition, analysis of data. P.A., M.P., K.N. and S.S. have contributed to the interpretation of data. P.A. and M.P. have drafted the work. V.P., H.M., J.B., M.D., P.M., R.C.B.-D., C.N., A.K., R.J., M.G., S.G.M., S.H., K.N. and S.S. substantively revised the manuscript.

All authors have approved the submitted version of the manuscript.

All authors have agreed both to be personally accountable for the author’s own contributions and to ensure that questions related to the accuracy or integrity of any part of the work, even ones in which the author was not personally involved, are appropriately investigated, resolved, and the resolution documented in the literature.

## Acknowledgements

Not applicable.

## References

1. Toledo JB, Arnold SE, Raible K, Brettschneider J, Xie SX, Grossman M, et al. Contribution of cerebrovascular disease in autopsy confirmed neurodegenerative disease cases in the National Alzheimer’s Coordinating Centre. Brain. 2013;136:2697–706. doi:10.1093/brain/awt188.

2. Schneider JA, Bennett DA. Where vascular meets neurodegenerative disease. Stroke. 2010;41:S144–6. doi:10.1161/STROKEAHA.110.598326.

3. Hainsworth AH, Markus HS, Schneider JA. Cerebral Small Vessel Disease, Hypertension, and Vascular Contributions to Cognitive Impairment and Dementia. Hypertension. 2024;81:75–86. doi:10.1161/HYPERTENSIONAHA.123.19943.

4. Yu L, Wang T, Wilson RS, Leurgans S, Schneider JA, Bennett DA, Boyle PA. Common age-related neuropathologies and yearly variability in cognition. Ann Clin Transl Neurol. 2019;6:2140–9. doi:10.1002/acn3.50857.

5. Wardlaw JM, Smith EE, Biessels GJ, Cordonnier C, Fazekas F, Frayne R, et al. Neuroimaging standards for research into small vessel disease and its contribution to ageing and neurodegeneration. Lancet Neurol. 2013;12:822–38. doi:10.1016/S1474-4422(13)70124-8.

6. Jack CR, Albert MS, Knopman DS, McKhann GM, Sperling RA, Carrillo MC, et al. Introduction to the recommendations from the National Institute on Aging-Alzheimer’s Association workgroups on diagnostic guidelines for Alzheimer’s disease. Alzheimers Dement. 2011;7:257–62. doi:10.1016/j.jalz.2011.03.004.

7. Song I-U, Kim J-S, Kim Y-I, Eah K-Y, Lee K-S. Clinical significance of silent cerebral infarctions in patients with Alzheimer disease. Cogn Behav Neurol. 2007;20:93–8. doi:10.1097/wnn.0b013e31805d859e.

8. Waziry R, Chibnik LB, Bos D, Ikram MK, Hofman A. Risk of hemorrhagic and ischemic stroke in patients with Alzheimer disease: A synthesis of the literature. Neurology. 2020;94:265–72. doi:10.1212/WNL.0000000000008924.

9. Lao PJ, Brickman AM. Multimodal neuroimaging study of cerebrovascular disease, amyloid deposition, and neurodegeneration in Alzheimer’s disease progression. Alzheimers Dement (Amst). 2018;10:638–46. doi:10.1016/j.dadm.2018.08.007.

10. Garnier-Crussard A, Bougacha S, Wirth M, Dautricourt S, Sherif S, Landeau B, et al. White matter hyperintensity topography in Alzheimer’s disease and links to cognition. Alzheimers Dement. 2022;18:422–33. doi:10.1002/alz.12410.

11. Benedictus MR, Prins ND, Goos JDC, Scheltens P, Barkhof F, van der Flier WM. Microbleeds, Mortality, and Stroke in Alzheimer Disease: The MISTRAL Study. JAMA Neurol. 2015;72:539–45. doi:10.1001/jamaneurol.2015.14.

12. Chin KS, Holper S, Loveland PM, Churilov L, Yassi N, Watson R. Prevalence of cerebral microbleeds in Alzheimer’s disease, dementia with Lewy bodies and Parkinson’s disease dementia: a systematic review and meta-analysis. Alzheimer’s & Dementia 2023. doi:10.1002/alz.079362.

13. Raposo N, Planton M, Péran P, Payoux P, Bonneville F, Lyoubi A, et al. Florbetapir imaging in cerebral amyloid angiopathy-related hemorrhages. Neurology. 2017;89:697–704. doi:10.1212/WNL.0000000000004228.

14. Jung YH, Lee H, Kim HJ, Na DL, Han HJ, Jang H, Seo SW. Prediction of amyloid β PET positivity using machine learning in patients with suspected cerebral amyloid angiopathy markers. Sci Rep. 2020;10:18806. doi:10.1038/s41598-020-75664-8.

15. Jung YH, Jang H, Park SB, Choe YS, Park Y, Kang SH, et al. Strictly Lobar Microbleeds Reflect Amyloid Angiopathy Regardless of Cerebral and Cerebellar Compartments. Stroke. 2020;51:3600–7. doi:10.1161/STROKEAHA.119.028487.

16. Jang H, Chun MY, Kim HJ, Na DL, Seo SW. The effects of imaging markers on clinical trajectory in cerebral amyloid angiopathy: a longitudinal study in a memory clinic. Alzheimers Res Ther. 2023;15:14. doi:10.1186/s13195-023-01161-5.

17. Charidimou A, Boulouis G, Frosch MP, Baron J-C, Pasi M, Albucher JF, et al. The Boston criteria version 2.0 for cerebral amyloid angiopathy: a multicentre, retrospective, MRI-neuropathology diagnostic accuracy study. Lancet Neurol. 2022;21:714–25. doi:10.1016/S1474-4422(22)00208-3.

18. McKhann G, Drachman D, Folstein M, Katzman R, Price D, Stadlan EM. Clinical diagnosis of Alzheimer’s disease: report of the NINCDS-ADRDA Work Group under the auspices of Department of Health and Human Services Task Force on Alzheimer’s Disease. Neurology. 1984;34:939–44. doi:10.1212/WNL.34.7.939.

19. Jack CR, Bennett DA, Blennow K, Carrillo MC, Dunn B, Haeberlein SB, et al. NIA-AA Research Framework: Toward a biological definition of Alzheimer’s disease. Alzheimer’s & Dementia. 2018;14:535–62. doi:10.1016/j.jalz.2018.02.018.

20. Wahlund LO, Barkhof F, Fazekas F, Bronge L, Augustin M, Sjögren M, et al. A new rating scale for age-related white matter changes applicable to MRI and CT. Stroke. 2001;32:1318–22. doi:10.1161/01.str.32.6.1318.

21. Jang H, Kim HJ, Park S, Park YH, Choe Y, Cho H, et al. Application of an amyloid and tau classification system in subcortical vascular cognitive impairment patients. Eur J Nucl Med Mol Imaging. 2020;47:292–303. doi:10.1007/s00259-019-04498-y.

22. Tsai Y-C, Tsai H-H, Liu C-J, Lin S-S, Chen Y-F, Jeng J-S, et al. Cerebral amyloid deposition predicts long-term cognitive decline in hemorrhagic small vessel disease. Brain Behav. 2023;13:e3189. doi:10.1002/brb3.3189.

23. Jang H, Jang YK, Kim HJ, Werring DJ, Lee JS, Choe YS, et al. Clinical significance of amyloid β positivity in patients with probable cerebral amyloid angiopathy markers. Eur J Nucl Med Mol Imaging. 2019;46:1287–98. doi:10.1007/s00259-019-04314-7.

24. Koemans EA, Chhatwal JP, van Veluw SJ, van Etten ES, van Osch MJP, van Walderveen MAA, et al. Progression of cerebral amyloid angiopathy: a pathophysiological framework. Lancet Neurol. 2023;22:632–42. doi:10.1016/S1474-4422(23)00114-X.

25. Spina S, La Joie R, Petersen C, Nolan AL, Cuevas D, Cosme C, et al. Comorbid neuropathological diagnoses in early versus late-onset Alzheimer’s disease. Brain. 2021;144:2186–98. doi:10.1093/brain/awab099.

26. Coomans EM, van Westen D, Binette AP, Strandberg O, Spotorno N, Serrano GE, et al. Interactions between vascular burden and amyloid-β pathology on trajectories of tau accumulation. Brain. 2024;147:949–60. doi:10.1093/brain/awad317.

27. Power MC, Mormino E, Soldan A, James BD, Yu L, Armstrong NM, et al. Combined neuropathological pathways account for age-related risk of dementia. Ann Neurol. 2018;84:10–22. doi:10.1002/ana.25246.

28. Leijenaar JF, Groot C, Sudre CH, Bergeron D, Leeuwis AE, Cardoso MJ, et al. Comorbid amyloid-β pathology affects clinical and imaging features in VCD. Alzheimer’s & Dementia. 2020;16:354–64. doi:10.1016/j.jalz.2019.08.190.

29. Kuo P-Y, Tsai H-H, Lee B-C, Chiang P-T, Liu C-J, Chen Y-F, et al. Differences in lobar microbleed topography in cerebral amyloid angiopathy and hypertensive arteriopathy. Sci Rep. 2024;14:3774. doi:10.1038/s41598-024-54243-1.

30. Pasi M, Charidimou A, Boulouis G, Auriel E, Ayres A, Schwab KM, et al. Mixed-location cerebral hemorrhage/microbleeds: Underlying microangiopathy and recurrence risk. Neurology. 2018;90:e119–e126. doi:10.1212/WNL.0000000000004797.

31. Jack CR, Knopman DS, Chételat G, Dickson D, Fagan AM, Frisoni GB, et al. Suspected non-Alzheimer disease pathophysiology--concept and controversy. Nat Rev Neurol. 2016;12:117–24. doi:10.1038/nrneurol.2015.251.

32. Kern S, Syrjanen JA, Blennow K, Zetterberg H, Skoog I, Waern M, et al. Association of Cerebrospinal Fluid Neurofilament Light Protein With Risk of Mild Cognitive Impairment Among Individuals Without Cognitive Impairment. JAMA Neurol. 2019;76:187–93. doi:10.1001/jamaneurol.2018.3459.

33. Cousins KAQ, Phillips JS, Irwin DJ, Lee EB, Wolk DA, Shaw LM, et al. ATN incorporating cerebrospinal fluid neurofilament light chain detects frontotemporal lobar degeneration. Alzheimer’s & Dementia. 2021;17:822–30. doi:10.1002/alz.12233.

34. Assmann A, Scheumann V, Ludwig AM, Garz C, Perosa V, Schreiber F, et al. P98. CSF NFL – A new biomarker for neurodegeneration in CSVD? Clinical Neurophysiology. 2018;129:e103–e104. doi:10.1016/j.clinph.2018.04.720.

35. Qu Y, Tan C-C, Shen X-N, Li H-Q, Cui M, Tan L, et al. Association of Plasma Neurofilament Light With Small Vessel Disease Burden in Nondemented Elderly: A Longitudinal Study. Stroke. 2021;52:896–904. doi:10.1161/STROKEAHA.120.030302.

36. Meeker KL, Butt OH, Gordon BA, Fagan AM, Schindler SE, Morris JC, et al. Cerebrospinal fluid neurofilament light chain is a marker of aging and white matter damage. Neurobiol Dis. 2022;166:105662. doi:10.1016/j.nbd.2022.105662.

37. Peters N, van Leijsen E, Tuladhar AM, Barro C, Konieczny MJ, Ewers M, et al. Serum Neurofilament Light Chain Is Associated with Incident Lacunes in Progressive Cerebral Small Vessel Disease. J Stroke. 2020;22:369–76. doi:10.5853/jos.2019.02845.

38. Wilcock D, Jicha G, Blacker D, Albert MS, D’Orazio LM, Elahi FM, et al. MarkVCID cerebral small vessel consortium: I. Enrollment, clinical, fluid protocols. Alzheimer’s & Dementia. 2021;17:704–15. doi:10.1002/alz.12215.

39. Iida MA, Farrell K, Walker JM, Richardson TE, Marx GA, Bryce CH, et al. Predictors of cognitive impairment in primary age-related tauopathy: an autopsy study. Acta Neuropathol Commun. 2021;9:134. doi:10.1186/s40478-021-01233-3.

40. Crary JF, Trojanowski JQ, Schneider JA, Abisambra JF, Abner EL, Alafuzoff I, et al. Primary age-related tauopathy (PART): a common pathology associated with human aging. Acta Neuropathol. 2014;128:755–66. doi:10.1007/s00401-014-1349-0.

41. Bennett RE, Robbins AB, Hu M, Cao X, Betensky RA, Clark T, et al. Tau induces blood vessel abnormalities and angiogenesis-related gene expression in P301L transgenic mice and human Alzheimer’s disease. Proc Natl Acad Sci U S A. 2018;115:E1289–E1298. doi:10.1073/pnas.1710329115.

42. Ye S, Pan H, Li W, Wang B, Xing J, Xu L. High serum amyloid A predicts risk of cognitive impairment after lacunar infarction: Development and validation of a nomogram. Front Neurol. 2022;13:972771. doi:10.3389/fneur.2022.972771.

43. Baiardi S, Abu-Rumeileh S, Rossi M, Zenesini C, Bartoletti-Stella A, Polischi B, et al. Antemortem CSF Aβ42/Aβ40 ratio predicts Alzheimer’s disease pathology better than Aβ42 in rapidly progressive dementias. Ann Clin Transl Neurol. 2019;6:263–73. doi:10.1002/acn3.697.

44. Rizzi L, Balthazar MLF. Mini-review: The suspected non-Alzheimer’s disease pathophysiology. Neurosci Lett. 2021;764:136208. doi:10.1016/j.neulet.2021.136208.

