## Supplementary file for "Alzheimer’s disease and neurodegeneration in symptomatic cerebral small vessel disease"

### Supplemental File

### Supplementary Material

#### Material and Methods

*Clinical data and neuropsychological assessment*

Vascular risk factors were defined by prior diagnosis of arterial hypertension, dyslipidemia and type 2 diabetes or antihypertensive, lipid lowering or antidiabetic medication. Additionally, we considered clinical laboratory blood tests for dyslipidemia (total cholesterol > 5.2 mmol/L, low density lipoprotein cholesterol > 2.6 mmol/L, high density lipoprotein cholesterol < 1.0 mmol/L or triglycerides > 1.7 mmol/L) and type 2 diabetes (HbA1c ≥ 6.5% or fasting plasma glucose level ≥ 7.0 mmol/L). The median time interval between the different diagnostic measures was as follows: MRI and LP 6 days (IQR 1 - 77); MRI and MMSE 41 days (IQR 4 - 188); LP and MMSE 15 days (IQR 2-180). In CSVD patients with previous symptomatic stroke, timespan between symptom onset and MMSE was 137 days (IQR 12-1609).

*Cerebrospinal fluid*

CSF samples were centrifuged at 4 °C, aliquoted and stored at -80 °C until analysis. Biomarker levels were determined with ELISA Kits (until 12/2019: Innotest $A\beta_{40}$, Innotest $A\beta_{42}$, Innotest pTau, Innotest hTauAg, Innogenetics, Ghent, Belgium; NfL Umandiagnostics, Sweden) or automated immunoassays (LUMIPULSE® G600 II, Fujirebio Inc., Japan, from 01/2020). Locally established thresholds were as follows: 0.50 for $A\beta_{42/40}$ ratio, 70 pg/mL for pTau, 350 pg/mL for tTau, 3643 pg/mL for NfL using ELISA kits and 0.69 for $A\beta_{42/40}$ ratio, 56 pg/mL for pTau, 404 pg/mL for tTau using immunoassays.[1,2] In line with ATN, each participant was classified as normal (−) or abnormal (+) for A based on the $A\beta_{42/40}$ ratio ([$A\beta_{42}$ / $A\beta_{40}$] x 10), for T based on pTau and for N based on tTau or NfL.

*MRI aquisition and analysis*

MRI was performed using a 3T (Siemens Heathineers, Erlangen, Germany; n = 92, 40%; n = 12 controls, n = 30 DPA, n = 37 CAA, n = 14 AD) or 1.5T MRI (Siemens Heathineers, Erlangen, Germany; n = 137, 60%; n = 6 controls, n = 32 DPA, n = 42 CAA, n = 57 AD). The following sequences were used to quantify CSVD markers according to Standards for Reporting Vascular Changes on Neuroimaging (STRIVE) consensus criteria[3]: T2*-weighted gradient-recalled echo for CMB and intracerebral hemorrhage (ICH) and T2-weighted fluid-attenuated inversion recovery for white matter hyperintensities

(WMH) and lacunes. MRI analysis of all participants was performed by one trained investigator (MP), blinded to demographic and clinical information. The images were evaluated using Mango software for dicom images (https://ric.uthscsa.edu/mango/). Presence and frequency of deep or lobar CMB, ICH or lacunes were assessed by applying the the Microbleed Anatomomic Rating Scale (CMB and lacunes) and the Cerebral Haemorrhage Anatomical RaTing instrument.[4,5] WMH in deep and periventricular regions were rated according to the Fazekas scale.[6] Based on a sample of 23 randomly chosen cases across all diagnostic groups the intra- and inter-rater reliability (by ACJ, a second independent and blinded rater) was excellent for all investigated variables (>0.99 for intra- and >0.79 for inter-rater reliability).

### Supplementary Tables

**Supplementary Table 1. Neuroimaging features of CSVD in all clinical subgroups.**

| | **CON**<br>n = 18 | **DPA**<br>n = 62 | **CAA**<br>n = 79 | **AD**<br>n = 70 |
|---|---|---|---|---|
| **Hemorrhagic lesions** | | | | |
| Lobar ICH on MRI | 0 (0%) | 9 (15%) | 21 (27%) | 0 (0%) |
| Deep ICH on MRI | 0 (0%) | 2 (3%) | 0 (0%) | 0 (0%) |
| Number of CMB | 0 (0-0) | 11 (5-28) | 4 (2-25) | 0 (0-0) |
| Number of lobar CMB | 0 (0-0) | 7 (3-20) | 4 (2-24) | 0 (0-0) |
| Presence of lobar CMB | 0 (0%) | 56 (90%) | 79 (100%) | 11 (16%) |
| Number of deep CMB | 0 (0-0) | 3 (1-5) | 0 (0-0) | 0 (0-0) |
| Presence of deep CMB | 0 (0%) | 62 (100%) | 0 (0%) | 2 (3%) |
| **Non-hemorrhagic lesions** | | | | |
| Number of lacunes | 0 (0-0) | 1 (0-4) | 0 (0-1) | 0 (0-0) |
| Lacune presence | 0 (0%) | 35 (57%) | 29 (37%) | 11 (16%) |
| Lobar lacune presence | 0 (0%) | 24 (39%) | 25 (32%) | 8 (11%) |
| Deep lacune presence | 0 (0%) | 23 (37%) | 14 (18%) | 5 (7%) |
| Periventricular WMH | 1 (1-1) | 3 (2-3) | 3 (2-3) | 2 (1-3) |
| Deep WMH | 1 (1-1) | 3 (2-3) | 2 (2-3) | 1 (1-2) |

Abbreviations: AD, Alzheimer's disease; CAA, cerebral amyloid angiopathy; CMB, cerebral microbleeds; CON, controls; CSVD, cerebral small vessel disease; DPA, deep perforator arteriopathy; ICH, intracerebral hemorrhage; MRI, magnetic resonance imaging; WMH, white matter hyperintensities.

**Supplementary Table 2. ATN classification profiles within the AD pathology continuum in CSVD patients.**

| | **DPA**<br>n = 62 | **CAA**<br>n = 79 | **Univariate analysis** |
|---|---|---|---|
| A+T-N- | 5 (8%) | 5 (6%) | $\chi2 = 0.16$<br>$p = .690$ |
| A+T-N+ | 2 (3%) | 4 (5%) | $\chi2 = 0.29$<br>$p = .592$ |
| A+T+N- | 0 (0%) | 0 (0%) | |
| A+T+N+ | 2 (3%) | 21 (27%) | $\chi2 = 13.88$<br>**$p < .001$** |

Abbreviations: AD, Alzheimer's disease; CAA, cerebral amyloid angiopathy; CSVD, cerebral small vessel disease; DPA, deep perforator arteriopathy.

**Supplementary Table 3. Inter-group comparison of continuous biomarker levels.**

| | **CON** n = 18 | **DPA** n = 62 | **CAA** n = 79 | **AD dementia** n = 70 | **Univariate analysis** | **Significant pairwise post-hoc tests** |
|---|---|---|---|---|---|---|
| **AD pathology biomarker levels** | | | | | | |
| $A\beta_{42/40}$ ratio | 1.30 (1.15-1.52) | 0.94 (0.63-1.17) | 0.60 (0.45-0.92) | 0.41 (0.33-0.50) | H = 103.6 **p < .001** | CON, DPA, CAA vs. AD: **p < .001** CON, DPA vs. CAA: **p < .01** |
| pTau [pg/mL] | 42 (24-49) | 45 (30-60) | 66 (42-85) | 101 (82-136) | H = 102.2 **p < .001** | CON, DPA, CAA vs. AD: **p < .001** CON, DPA vs. CAA: **p < .01** |
| **Neurodegeneration biomarker levels** | | | | | | |
| tTau [pg/mL] | 182 (90-251) | 289 (175-460) | 420 (287-603) | 610 (493-830) | H = 82.17 **p < .001** | CON, DPA, CAA vs. AD: **p < .001** CON, DPA vs. CAA: **p < .02** CON vs. DPA: **p = .02** |
| NfL [ng/mL] | 1.14 (0.94-1.38) | 4.18 (2.54-9.68) | 3.67 (2.09-12.75) | 1.64 (1.35-2.19) | H = 50.12 **p < .001** | DPA, CAA vs. CON, AD: **p < .001** |

Abbreviations: Aβ, amyloid-beta; AD, Alzheimer's disease; CAA, cerebral amyloid angiopathy; CON, controls; DPA, deep perforator arteriopathy; NfL, Neurofilament light; pTau, phosphorylated-tau; tTau, total-tau.

**Supplementary Table 4. CSVD patient characteristics associated with pathological ATN profiles.**

| | **A+T±N±**<br>n = 39 | **Not A+T±N±**<br>n = 102 | p-value | **A-T+N+**<br>n = 18 | **Not A-T+N+**<br>n = 123 | p-value | **A-T-N+**<br>n = 41 | **Not A-T-N+**<br>n = 100 | p-value |
|---|---|---|---|---|---|---|---|---|---|
| Age | 77 (73-81) | 72 (65-77) | **<.001** | 77 (74-80) | 73 (65-78) | **.047** | 69 (62-76) | 75 (68-80) | **<.001** |
| Female sex | 22 (56%) | 37 (38%) | **.049** | 8 (44%) | 51 (43%) | .914 | 13 (32%) | 48 (48%) | .076 |
| Cognitive impairment | 27 (69%) | 53 (52%) | .064 | 13 (72%) | 67 (54%) | .156 | 20 (49%) | 60 (60%) | .222 |
| Previous stroke | 14 (36%) | 41 (40%) | .640 | 5 (28%) | 50 (41%) | .296 | 25 (61%) | 30 (30%) | **< .001** |
| Previous seizure | 8 (21%) | 22 (22%) | .891 | 4 (22%) | 26 (21%) | .916 | 10 (24%) | 20 (20%) | .563 |
| Gait disturbances | 8 (21%) | 26 (26%) | .537 | 3 (17%) | 31 (25%) | .429 | 13 (32%) | 21 (21%) | .177 |
| Presence of lobar ICH | 14 (36%) | 18 (18%) | **.021** | 3 (17%) | 29 (24%) | .513 | 9 (22%) | 23 (23%) | .893 |
| Number of total CMB | 16 (3-47) | 7 (3-18) | **.032** | 5 (1-9) | 10 (3-28) | .061 | 15 (4-27) | 7 (3-25) | .095 |
| Number of lobar CMB | 10 (3-47) | 5 (2-13) | **.011** | 5 (1-7) | 6 (2-25) | .114 | 9 (2-23) | 5 (2-21) | .438 |
| Number of deep CMB | 0 (0-0) | 1 (0-3) | **.008** | 0 (0-1) | 0 (0-3) | .280 | 2 (0-5) | 0 (0-1) | **<.001** |
| Number of lacunes | 0 (0-1) | 1 (0-3) | **.002** | 0 (0-4) | 0 (0-2) | .879 | 1 (0-4) | 0 (0-2) | **.015** |
| Presence of lobar lacune | 8 (21%) | 40 (39%) | **.036** | 7 (39%) | 41 (33%) | .642 | 17 (42%) | 31 (31%) | .234 |
| Presence of deep lacune | 3 (8%) | 34 (33%) | **.002** | 5 (28%) | 32 (26%) | .874 | 15 (37%) | 22 (22%) | .074 |
| Periventricular WMH | 2 (2-3) | 3 (2-3) | .220 | 3 (2-3) | 3 (2-3) | .186 | 3 (2-3) | 3 (2-3) | .414 |
| Deep WMH | 2 (1-3) | 3 (2-3) | **.036** | 3 (2-3) | 2 (2-3) | .065 | 3 (2-3) | 2 (2-3) | .079 |

Abbreviations: CMB, cerebral microbleeds; CSVD, cerebral small vessel disease; ICH, intracerebral hemorrhage; WMH, white matter hyperintensities.

**Supplementary Table 5. Predictors of pathological ATN profiles in DPA and CAA.**

| | DPA | | | CAA | | |
|---|---|---|---|---|---|---|
| | **A+T±N±**<br>n = 9 | **A-T±N±**<br>n = 53 | **p-value** | **A+T±N±**<br>n = 30 | **A-T±N±**<br>n = 49 | **p-value** |
| Age | 80 (74-83) | 69 (60-77) | **.004** | 77 (73-81) | 75 (69-79) | .175 |
| Female sex | 8 (89%) | 19 (36%) | **.003** | 14 (47%) | 20 (41%) | .610 |
| Presence of lobar ICH | 1 (11%) | 9 (17%) | .658 | 13 (43%) | 9 (18%) | **.016** |
| Number of lobar CMB | 7 (2-60) | 7 (3-18) | .421 | 16 (3-47) | 3 (1-9) | **.006** |
| Presence of deep lacune | 1 (11%) | 22 (42%) | .081 | 2 (7%) | 12 (25%) | **.044** |
| Deep WMH | 2 (1-3) | 3 (2-3) | .072 | 2 (2-3) | 3 (2-3) | .347 |
| | **A-T+N+**<br>n = 6 | **Not A-T+N+**<br>n = 56 | **p-value** | **A-T+N+**<br>n = 12 | **Not A-T+N+**<br>n = 67 | **p-value** |
| Age | 76 (72-83) | 70 (60-78) | .076 | 78 (69-80) | 75 (70-80) | .511 |
| | **A-T-N+**<br>n = 26 | **Not A-T-N+**<br>n = 36 | **p-value** | **A-T-N+**<br>n = 15 | **Not A-T-N+**<br>n = 64 | **p-value** |
| Age | 64 (60-75) | 75 (65-81) | **.025** | 72 (69-76) | 75 (70-80) | .213 |
| Previous symptomatic stroke | 16 (62%) | 10 (28%) | **.008** | 9 (60%) | 20 (31%) | **.038** |
| Number of lacunes | 2 (0-6) | 1 (0-4) | .154 | 0 (0-3) | 0 (0-1) | .258 |

Abbreviations: CAA, cerebral amyloid angiopathy; CMB, cerebral microbleeds; DPA, deep perforator arteriopathy; ICH, intracerebral hemorrhage; WMH, white matter hyperintensities

**Supplementary Table 6. ATN profile distribution according to version of Boston criteria.**

| | **Probable CAA**<br>**Boston v1.5**<br>n = 51 | **Probable CAA**<br>**only Boston v2.0**<br>n = 28 | **p-value** |
|---|---|---|---|
| **A+T±N±** | 26 (51%) | 4 (14%) | **.001** |
| **A-T+N+** | 7 (14%) | 5 (18%) | .625 |
| **A-T-N+** | 9 (18%) | 6 (21%) | .682 |

Abbreviations: CAA, cerebral amyloid angiopathy.

### Supplementary Figure

**Supplementary Figure 1. Group comparison of continuous CSF biomarker levels.**

Turkey box plot diagrams display (**A**) amyloid, (**B**) phosphorylated Tau and (**C**, **D**) neurodegenerative CSF biomarkers. NfL measurements were available for n = 139 (59%) patients only. *p < .05, **p < .01, ***p < .001, ****p < .0001.

Abbreviations: Aβ, amyloid-beta; AD, Alzheimer's disease dementia; CAA, cerebral amyloid angiopathy; CON, controls; CSF, cerebrospinal fluid; DPA, deep perforator arteriopathy; NfL, Neurofilament L; pTau, phosphorylated-tau; tTau, total-tau.

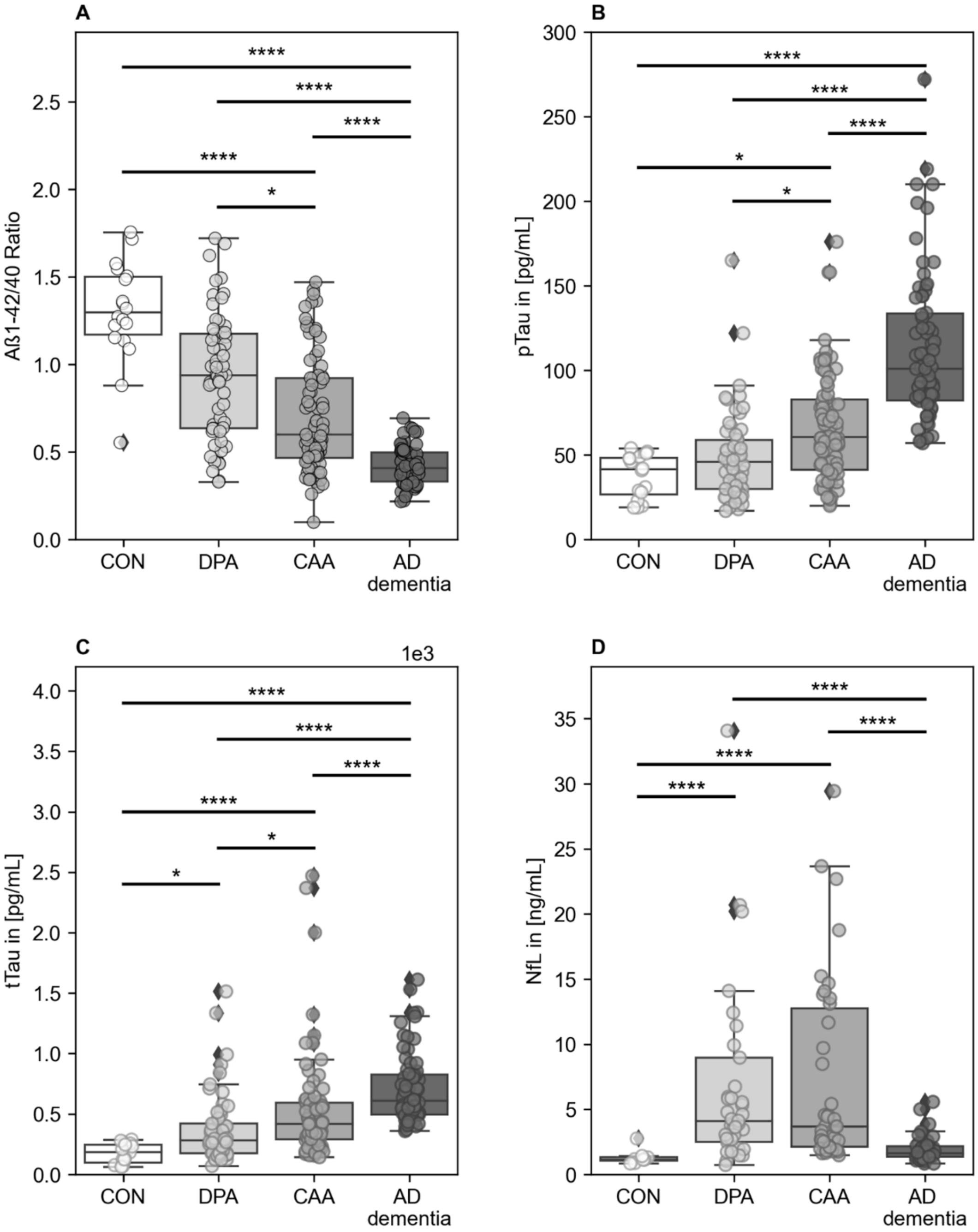

### References

1. Perosa V, Priester A, Ziegler G, et al. Hippocampal vascular reserve associated with cognitive performance and hippocampal volume. *Brain*. 2020;143(2):622-634. doi:10.1093/brain/awz383.
2. Körtvelyessy P, Heinze HJ, Prudlo J, Bittner D. CSF Biomarkers of Neurodegeneration in Progressive Non-fluent Aphasia and Other Forms of Frontotemporal Dementia: Clues for Pathomechanisms? *Front Neurol*. 2018;9:504. doi:10.3389/fneur.2018.00504.
3. Wardlaw JM, Smith EE, Biessels GJ, et al. Neuroimaging standards for research into small vessel disease and its contribution to ageing and neurodegeneration. *Lancet Neurol*. 2013;12(8):822-838. doi:10.1016/S1474-4422(13)70124-8.
4. Gregoire SM, Chaudhary UJ, Brown MM, et al. The Microbleed Anatomical Rating Scale (MARS): reliability of a tool to map brain microbleeds. *Neurology*. 2009;73(21):1759-1766. doi:10.1212/WNL.0b013e3181c34a7d.
5. Charidimou A, Schmitt A, Wilson D, et al. The Cerebral Haemorrhage Anatomical RaTing inStrument (CHARTS): Development and assessment of reliability. *J Neurol Sci*. 2017;372:178-183. doi:10.1016/j.jns.2016.11.021.
6. Wahlund LO, Barkhof F, Fazekas F, et al. A new rating scale for age-related white matter changes applicable to MRI and CT. *Stroke*. 2001;32(6):1318-1322. doi:10.1161/01.str.32.6.1318.
